# Comparison of horizontal corneal diameter measurements using Orbscan IIz, OPD Scan III, and IOLMaster 700

**DOI:** 10.1101/2020.05.21.20109488

**Authors:** Sebastian Cruz, Felipe Valenzuela, Juan Stoppel, Eugenio Maul, Allister Gibbons

## Abstract

**Purpose:** To compare 3 automated devices for measuring the horizontal corneal diameter [white-to-white (WTW) distance].

**Setting:** Fundacion Oftalmologica Los Andes, Santiago, Chile.

**Study Design:** Retrospective.

**Methods:** In 65 eyes of 38 patients, the WTW distance was measured independently using Orbscan IIz tomography system (Bausch & Lomb), IOLMaster 700 (Carl Zeiss Meditec) and OPD Scan III (NIDEK). We tested for systematic differences in measurements and estimated the limits of agreement (LoA) using linear mixed effects models.

**Results:** The mean WTW distance was 11.8 ± 0.40 mm with Orbscan IIz, 12.1 ± 0.5 mm with IOLMaster 700 and 12.0 ± 0.4 mm with OPD Scan III. The mean difference between IOLMaster 700 and Orbscan IIz was 0.33 (95% CI 0.28;0.38) (p<0.001), between OPD Scan III and Orbscan IIz was 0.24 mm (95% CI 0.21;0.28) (p<0.001), and between IOL Master 700 and OPD Scan III was 0.09 (95% CI 0.05;0.12) (p<0.001). The 95% LoA for Orbscan IIz versus IOLMaster 700 was −0.69 mm to 0.03 mm, Orbscan IIz versus OPD Scan III was −0.52 mm to −0.03 mm, and OPD versus IOLMaster 700 was −0.39 mm to 0.22 mm. Switching to IOLMaster 700 or OPD Scan III measurements led to a selection of a longer phakic IOL length (Visian ICL, STAAR) in 34% and 33% of the cases, respectively compared to Orbscan IIz.

**Conclusions:** The data suggests that these devices are not interchangeable for usual clinical practice. Adjustments based on mean differences was not enough to compensate for inter-instrument discrepancy in WTW measurements.

## INTRODUCTION

The horizontal corneal diameter, also known as white to white (WTW) distance is the measurement between the limbus from one side of the cornea to the other and is essential for sizing calculation of the Implantable Collamer Lens (ICL) posterior chamber phakic intraocular lens (pIOL) (STAAR Surgical, Monrovia, California). ^1-3^ Many postoperative complications associated with posterior chamber phakic IOL surgery are related to suboptimal lens sizing. ^4,5^ An oversized lens can cause angle closure or iris chafing leading to pigment dispersion, whereas an undersized lens can accelerate anterior subcapsular cataract formation or cause zonular damage with subsequent dislocation of the pIOL. ^4-7^

The lens sizing protocol originally approved by the United States Food and Drug Administration (FDA) requires adding 0.5mm to the to the horizontal white-to-white measurement obtained using calipers at a slit lamp or using the Orbscan unit (Bausch & Lomb, Rochester, NY). ^1^ There are manual and automated methods of WTW distance measurement. ^8-10^ Manual methods include millimeter rules, photography, calipers, and the direct beam of slit lamp in coaxial position. ^11,12^ Automated methods, like the ones compared in this study, use computerized video limbus detection through digital gray-scale and have been associated to reduced inter-operator variability and greater accuracy. ^11^

Comparison studies including tomographers such as the Orbscan IIz and Pentacam HR (Oculus, Wetzlar, Germany) have determined a high level of correlation, and some authors have even suggested that the data can be used interchangeably in clinical practice.^13^ However, studies comparing other instruments: IOLMaster 500 (Carl Zeiss Meditec, Jena, Germany), LenStar 900 (HaagStreit, Wedel, Germany), Pentacam HR, Visante OCT (Carl Zeiss Meditec) have shown that measurements are not interchangeable. ^14^ The IOLMaster 700 (Carl Zeiss Meditec AG, Jena, Germany), which was the first commercially available swept source optical coherence tomography (SS-OCT) based biometer, was introduced in 2015 in the USA. To the best of our knowledge there are no studies comparing the WTW distance between IOL Master 700 and Orbscan IIz, neither between the OPD Scan III aberrometer (Nidek, Gamagori, Japan) versus other automated instruments.

The purpose of this study was to compare automated horizontal corneal diameter (WTW distance) measurements using the IOLMaster 700, OPD Scan III and Orbscan IIz and to determine the agreement between them.

## SUBJECTS AND METHODS

### Patients

This study consisted of a single-center retrospective review of patients who were evaluated for refractive surgery with 3 different automated WTW measurements: Orbscan IIz, OPD Scan III and IOL Master 700. The measurements were performed independently by 3 examiners on the same visit day. This study was conducted at Fundación Oftalmológica Los Andes (Santiago, Chile), this study was approved by our institution’s Ethics Committee and adhered to the tenets of the Declaration of Helsinki. Patients with known limbal pathology, previous ocular surgery, trauma or anatomical corneal alterations were excluded. Cases with evident failure of the automated segmentation algorithm for any of the 3 instruments were also excluded.

### Instruments

The Orbscan IIz device combines the Placido rings and slit-scanning method to improve the precision of topographic measurements. The device provides data on corneal topography, corneal thickness, and anterior chamber depth. A digital gray-scale image of the anterior segment is reconstructed from 140 slit images. The computer automatically detects the cornea by comparing grayscale steps and calculates the horizontal corneal diameter. ^13^

The IOLMaster 700 is the first SS-OCT-based biometry that measures a longitudinal section of the eye (AL, anterior chamber depth, corneal central thickness, lens thickness) based on SS-OCT with the exception of WTW, which is measured using the light-emitting diode light source determined by iris configuration, and keratometry, which is measured using 2.5-m zone telecentric keratometry. For WTW measurements, the limbus is detected automatically by a digital gray-scale photograph of the anterior eye segment is taken after focusing on the iris. ^15,16^ The OPD Scan III is capable of detecting the limbus, in a similar way to Orbscan IIz. It allows the measurement of WTW distance in mesopic and scotopic conditions. The latter is an important aspect, considering that the measurement uses grayscale that is affected by the brightness. ^17^

### Measurements

Measurements were obtained, for each patient, during the same day. These were performed by 3 different certified and qualified operators that were not aware of the results obtained by the other 2 instruments. If necessary, imaging was repeated until exams with acceptable quality were obtained. If accurate images for any of these modalities were not obtained, for a given eye, that eye was excluded from the study. Furthermore, any patient whose values had to be manually adjusted were not included in the study. In the Orbscan IIz, the automatically detected edge of the cornea is displayed with a green broken line, although this equipment discards low quality images, in some cases, measurement errors related to limbus recognition can occur. For the measurement of IOLMaster 700, all the manufacturer’s instructions were followed. Measurements that were qualified as “Borderline value!” (uncertain value) were excluded. In OPD Scan III, the calculated corneal diameter is displayed in a similar way to the Orbscan IIz. To avoid bias and to standardize the conditions for all measurements, all tests were done under mesopic conditions.

### Statistical Analysis

Statistical analysis and plotting was performed using Stata version 13 (StataCorp College Station, Texas) and R 3.6 (www.r-project.com, Vienna Austria). To account for the correlation of measurements of patients in which right and left eyes were included, linear mixed effects models were used to test for systematic measurement differences between methods and estimate limits of agreement. Bland–Altman plots with 95% LoA were used to display inter-instrument measurement differences and limits of agreement. The McNemar test was used to compare de size miss-selection between IOLMaster 700 and OPD Scan III vs Orbscan IIz.

## RESULTS

Data from 78 eyes of 47 refractive surgery candidates were reviewed. A total of 13 eyes were excluded due to measurement error (failure of automated segmentation or unreliable exams), leaving 65 eyes of 38 patients for analysis. Ocular characteristics are shown in **Table 1**.

**Table 1.** Patients characteristics by laterality, axial length and anterior chamber depth.

|  | Number |
| --- | --- |
| **Eyes, n (%)** | 65 (100) |
| Right | 34 (52) |
| Left | 31 (48) |
| **AL mean, mm (SD)** | 26 (3) |
| **AL, n (%)** |  |
| <24 mm | 15 (23) |
| >26 mm | 35 (54) |
| 24-26 mm | 15 (23) |
| **ACD mean, mm (SD)** | 3.4 (0.4) |
| **ACD, n (%)** |  |
| <3.5mm | 37 (57) |
| >=3.5mm | 28 (43) |

SD= standard deviation, mm= millimeters, AL= axial length, ACD= anterior chamber depth

Excluded cases can be divided as follows: Orbscan IIz (4 eyes, 2 patients), OPD Scan III (7 eyes, 5 patients), IOLMaster 700 (2 eyes, 2 patients). The mean WTW distance measurement obtained with Orbscan IIz, OPD Scan III and IOLMaster 700 were 11.8 ± 0.4 mm, 12.0 ± 0.4 mm and 12.1 ± 0.5 mm, respectively. **Table 2** shows WTW measurements according to gender, laterality, axial length (AL) and anterior chamber depth (ACD).

**Table 2.** Mean WTW distance by gender, laterality, axial length and anterior chamber depth.

|  | IOL Master 700<br>mm (SD) | OPD Scan III<br>mm (SD) | Orbscan IIz<br>mm (SD) | p value | Total (n) |
| --- | --- | --- | --- | --- | --- |
| Overall | 12.1 (0.5) | 12.0 (0.4) | 11.8 (0.4) | 1.00E-04 | 65 |
| Right eye | 12.0 (0.5) | 12.0 (0.4) | 11.7 (0.4) | 0.0074 | 34 |
| Left eye | 12.1 (0.5) | 12.0 (0.5) | 11.8 (0.4) | 0.0118 | 31 |
| Male | 12.2 (0.4) | 12.1 (0.4) | 11.8 (0.4) | 0.0045 | 24 |
| Female | 12.0 (0.5) | 11.9 (0.4) | 11.7 (0.4) | 0.0121 | 41 |
| AL <24 mm | 12.0 (0.4) | 11.8 (0.4) | 11.7 (0.4) | 0.1564 | 15 |
| AL 24-26 mm | 12.3 (0.5) | 12.2 (0.4) | 11.9 (0.4) | 0.1008 | 15 |
| AL >26 mm | 12.1 (0.5) | 12.0 (0.4) | 11.7 (0.4) | 0.0037 | 35 |
| ACD <3.5 mm | 12.0 (0.4) | 11.9 (0.4) | 11.7 (0.4) | 0.0034 | 37 |
| ACD ≥3.5 mm | 12.3 (0.5) | 12.2 (0.5) | 11.9 (0.4) | 0.0091 | 28 |

SD= standard deviation, mm= millimeters, AL= axial length, ACD= anterior chamber depth

Figure 1 shows Bland-Altman plots for the 3 compared instruments. Inter-instrument differences do not vary significantly in magnitude or dispersion across the range of WTW values included.

The mean difference between IOLMaster 700 and Orbscan IIz was 0.33 (95% CI 0.28;0.38) (p<0.001). The mean difference between OPD Scan III and Orbscan IIz was 0.24 mm (95% CI 0.21;0.28) (p<0.001). All comparisons showed a statistically significant difference between these machines, with the IOLMaster 700 always measuring longer **(Table 3)**.

**Table 3.** Mean difference and 95% confidence interval in corneal diameter measurements for Orbscan IIz, OPD Scan III and IOLMaster 700.

| Measurement | Mean difference, mm (95% CI) | p value |
| --- | --- | --- |
| Orbscan-OPD | -0.24 (-0.21; -0.28) | <0.01 |
| Orbscan-IOL700 | -0.33 (-0.28; -0.38) | <0.01 |
| OPD-IOL700 | -0.09 (-0.05, -0.12) | <0.01 |

CI= confidence interval, mm: millimeters

The 95% limits of agreement for IOLMaster 700 vs Orbscan IIz (0.03 to −0.69mm) were wider than the other comparisons **(Figure 1 & Table 4)**.

**Figure 1.**
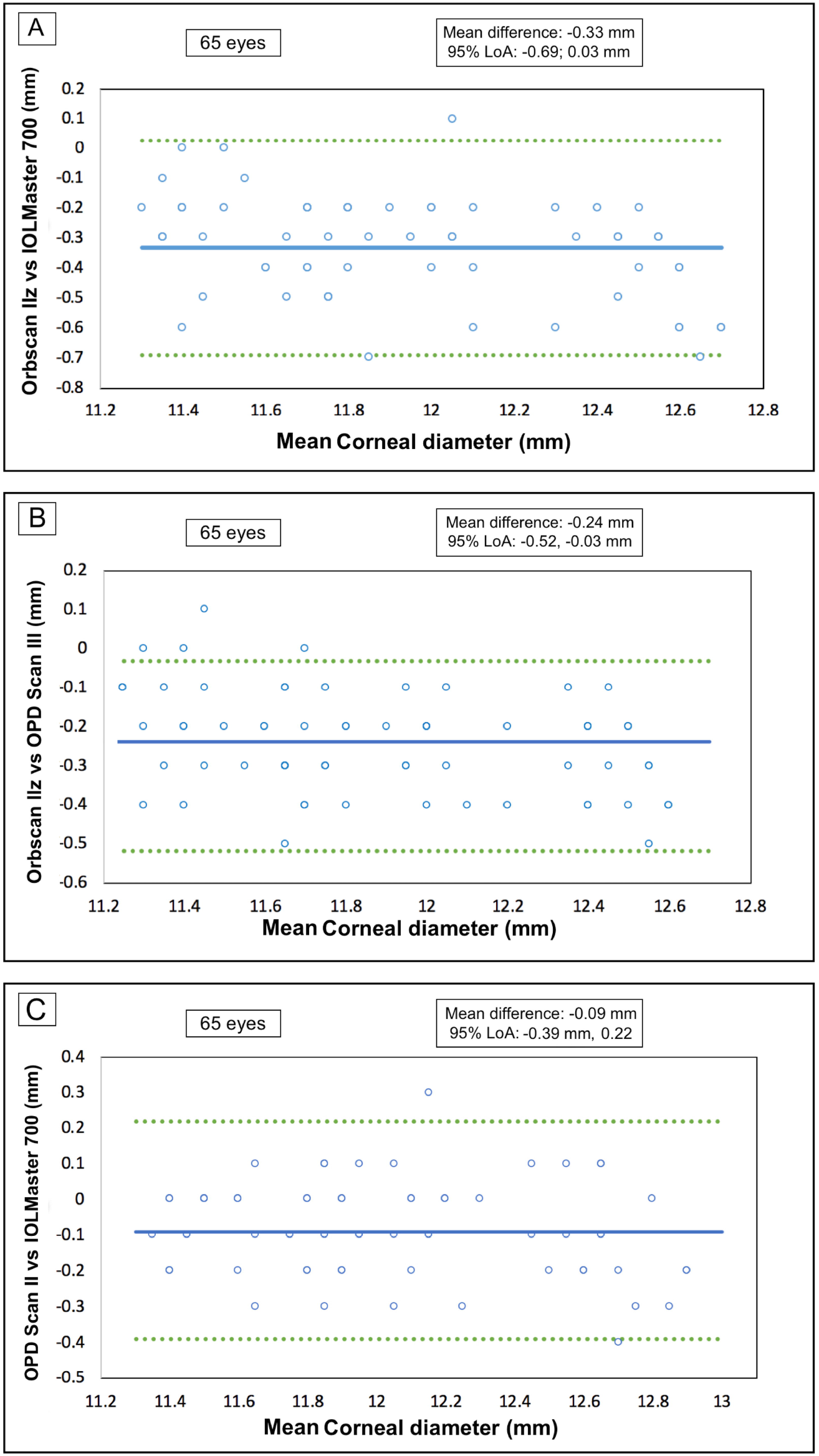

**Table 4.**
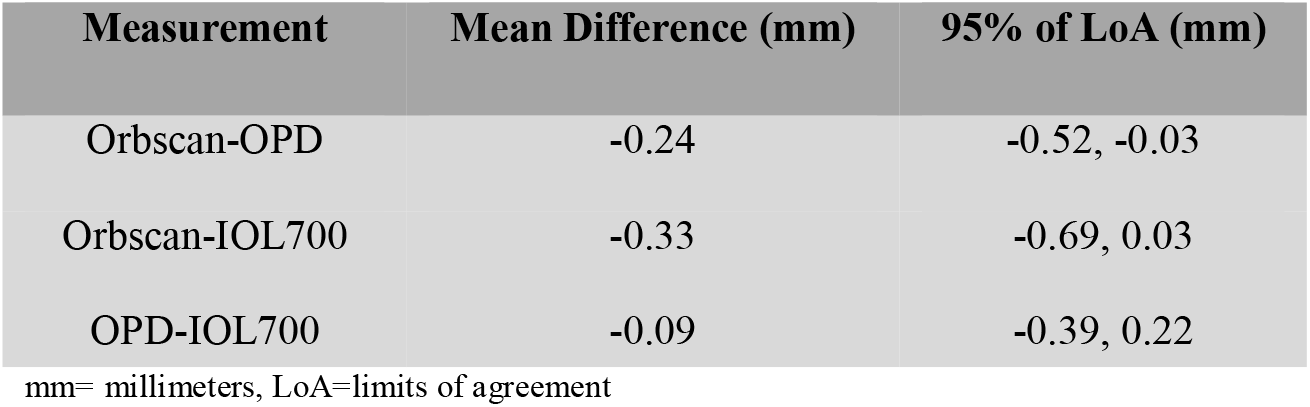
Mean difference and limits of agreement in corneal diameter measurements for Orbscan IIz, OPD Scan III and IOLMaster 700.

Using the STAAR surgical and ordering system OCOS™ website (https://evo-ocos.staarag.ch/Live/) we calculated ICL size based on Orbscan IIz, IOLMaster 700 and OPD III. Comparing the current recommended method of using WTW measurements obtained with Orbscan IIz to IOLMaster 700 WTW measurements, resulted in a longer lens length recommendation in 33.9% of the cases. Similarly, using the OPD Scan III WTW measurements resulted in a longer lens length recommendation in 32.3% of the cases.

Adjusting IOLMaster 700 and OPD Scan III WTW measurements, using the mean difference obtained in this study, led to a reduction in the discrepancy in calculated lens length. For IOLMaster 700, 21.5% of lenses had a different length compared to Orbscan IIz lens size estimation. For OPD Scan III, 20% of lenses had a different length compared to Orbscan IIz lens size estimation. Details of the lens length discrepancy before and after adjustment are depicted in Figure 2.

**Figure 2.**
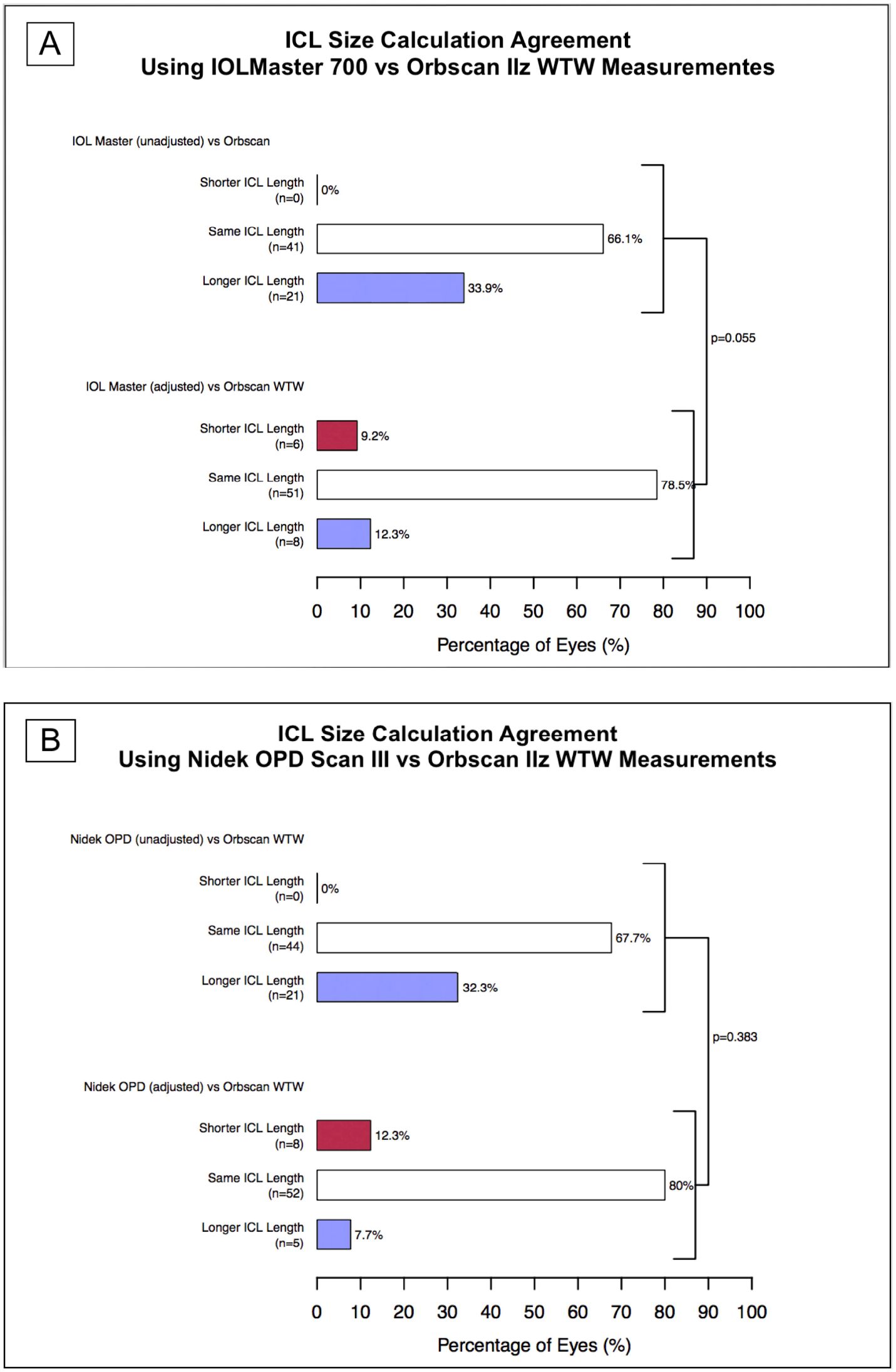

## DISCUSSION

The present study shows that the mean WTW distance readings with the IOLMaster 700 and the OPD Scan III are greater than those obtained with the Orbscan IIz, respectively. This difference was 0.33 mm and 0.24 mm in the case of the IOLMaster 700 and OPD Scan III, respectively. This would have had an impact in pIOL ICL selection, in over a 30% of cases, potentially leading to an increased rate of complications.

The manufacturer’s current recommended automated method for measuring the WTW in ICL candidates is based on Orbscan IIz.^1^ This has been studied extensively, and its acceptable accuracy, reproducibility and good clinical outcomes have already been reported. ^18^ To the best of our knowledge this is the first study comparing Orbscan IIz to the new swept source IOLMaster 700 and to OPD Scan III. According to our last search (MEDLINE browsing performed on April 30^th^, 2020), there were no publications regarding the interchangeability of WTW distance measurements obtained with the IOLMaster 700 or OPD Scan III and Orbscan IIz.

The 95% LoA for IOLMaster 700 versus Orbscan IIz obtained in our study were −0.69 mm to 0.03 mm, considerably wider than 95% LoA for Pentacam HR versus Orbscan IIz (−0.14 to 0.33 mm; mean difference 0.10 mm) reported by Salouti et al, but lower than Galilei versus Orbscan IIz (−0.72 to 1.48 mm, mean difference 0.38) reported previously by the same group. ^13,19^

In a previous publication by Baumeister et al using the IOLMaster 500 the mean WTW distance difference was approximately 0.24 mm longer with the IOLMaster 500 than with the Orbscan IIz. The 95% LoA reported for the IOLMaster 500 versus Orbscan IIz were −0.41 to 0.9 (LoA higher than our study). ^11^ Kohnen et al, showed a mean difference of 0.32 mm higher with the IOLMaster 500 versus Orbscan IIz, and 95% LoA between 0.1 to 0.54 mm (LoA which were lower than the ones found in our study). ^12^

Interestingly, in the present study OPD Scan III was the equipment that excluded the largest number of patients due to measurement errors. This suggests that it could have more difficulties in detecting the limbus compared to the other two methods. Therefore, we recommend carefully reviewing the readings obtained to be sure that the automatically detected edge of the cornea is displayed correctly.

According to our results, IOLMaster 700, OPD Scan III and Orbscan IIz cannot be used interchangeably, as the differences between them can be as great as 0.72 mm (Orbscan IIz/IOL Master 700), 0.55 mm (OPD Scan III/IOL Master 700) and 0.61 mm.\ (Orbscan IIz/OPD Scan III). Ophthalmologists who routinely use other automated instruments to measure WTW distance for posterior chamber pIOL implantations should consider existing differences between these instruments in their routine clinical practice.

ICL size selection is critical since it is directly related to postoperative vault which is defined as the distance between the anterior surface of the crystalline lens and the posterior surface of the ICL. This is crucial in avoiding adverse events; overvaulting, which can result in elevated intraocular pressure (IOP), glaucoma, endothelial cell loss, and pupil distortion; and low vaulting, which can accelerate cataract formation. ^4,5,7^All these complications can lead to the removal of the phakic lens, its replacement with an ICL of a more appropriate size, or even the need to render the patient pseudophakic.

One of the most commonly methods used to predict ICL postoperative vault is the WTW distance, which is used as a proxy for sulcus-to-sulcus (STS) distance. It has been proposed that direct STS measurement would be more accurate as there are studies that shown low to no correlation between WTW and STS dimensions. ^20,21^ However, methods to evaluate STS dimension are not widely available, are operator dependent and have not been conclusive to improve postoperative vault values. ^22^ With STS still not approved by the FDA and widespread non-availability of ultrasound biomicroscopy among refractive surgeons, the current search is towards instruments that can give the ideal WTW for estimating the ICL sizing with accuracy.

Dougherty et al and Kojima et al reported regression equations to calculate optimal ICL size.^23,24^ They used STS, ICL power, ACD and distance between STS plane and anterior crystalline lens surface as possible variables, leading to improved vaulting outcomes. ^23,24^ More studies are needed for evaluating accuracy and reliability of these models, which have not been validated by the manufacturer. Future refinement of these models may allow improvement in vault outcomes, thereby decreasing the adverse event rate in these patients.

There are several limitations in this study. First, all selected patients had normal corneas, so these results cannot be extrapolated to patients with corneal abnormalities. Second, this is a retrospective study and although those who performed the exams were qualified, only automated measurements with no errors were considered for evaluation.

In summary, WTW distance was on average shorter when measured with Orbscan IIz, followed by OPD Scan III and finally IOLMaster 700. However, our data does not support that raw or adjusted WTW measurements can be used interchangeably for pIOL ICL size calculation.

## Data Availability

What data in particular will be shared? Individual participant data that underlie the results reported in this article, after de-identification.
What documents will be available? Study protocol, statistical analysis plan. When will data be available? Immediately after publication (no end date) With whom? Researchers who provide a methodologically sound proposal
For what the of analyses? Any purpose
Proposals should be directed to Felipe Valenzuela, MD, Fundacion Oftalmologica los Andes.

